# Revisiting the effects of MDR1 Variants using computational approaches

**DOI:** 10.1101/2023.09.02.23294978

**Authors:** Tal Gutman, Tamir Tuller

## Abstract

P-glycoprotein, encoded by the MDR1 gene, is an ATP-dependent pump that exports various substances out of cells. Its overexpression is related to multi drug resistance in many cancers. Numerous studies explored the effects of MDR1 variants on p-glycoprotein expression and function, and on patient survivability. T1236C, T2677C and T3435C are prevalent MDR1 variants that are the most widely studied, typically in-vitro and in-vivo, with remarkably inconsistent results. In this paper we perform computational, data-driven analyses to assess the effects of these variants using a different approach. We use knowledge of gene expression regulation to elucidate the variants’ mechanism of action. Results indicate that T1236C increases MDR1 levels by 2-fold and is correlated with worse patient prognosis. Additionally, examination of MDR1 folding strength suggests that T3435C potentially modifies co-translational folding. Furthermore, all three variants reside in potential translation bottlenecks and likely cause increased translation rates. These results support several hypotheses suggested by previous studies. To the best of our knowledge, this study is the first to apply a computational approach to examine the effects of MDR1 variants.

## Introduction

P-glycoprotein (p-gp) was first discovered in 1976, where it was shown to alter drug permeability in Chinese hamster ovary cells^1^. Ensuing extensive studies unveiled ample information about the structure and function of this transmembrane protein; P-gp is a member of the ATP-binding cassette transporter superfamily^2^. It is a 170 KDa molecule that is comprised of two halves with high homology, each containing six transmembrane domains (TMD) and a single nucleotide-binding domain (NBD)^3^. P-gp is interleaved in the cell’s membrane and functions as an ATP-dependent efflux pump, exporting diverse substances out of the cell. Lipids, steroids, xenobiotics, various drugs and chemotherapy agents are some of the molecules transferred by this protein^4^. P-gp is normally highly expressed in the kidneys, liver, the gastro-intestinal tract and the blood-brain barrier^5^. It protects the body from deleterious substances by excreting them to the gut lumen, urine and bile, and by reducing permeability of sensitive tissues^6^. P-gp is also highly expressed in many cancer cells. Its ability to export substrates that vary greatly in chemical structure and size makes it fundamental for the multi-drug resistance (MDR) mechanism^7^ of cancer cells.

P-gp is encoded by the MDR1 gene, also frequently called ABCB1. It is located on chromosome 7 and encodes for a protein of 1,280 amino acids^8^. MDR1 is highly polymorphic, with over 50 single nucleotide polymorphisms (SNP) currently discovered in its coding region^9^. Three variants are the most extensively studied - T1236C, T2677G and T3435C. T1236C is a synonymous variant that occurs in the first NBD (at position 1236 of the coding sequence), changing the codon GGT to GGC (both encode for Glycine).

T2677G is a non-synonymous variant that occurs between the tenth and eleventh TMDs. It changes the codon TCT to GCT, resulting in Serine being replaced by Alanine. Finally, T3435C is a synonymous variant that occurs in the second NBD, causing the codon ATT to be changed to ATC but keeping the encoded amino acid Isoleucine^10^. All three variants occur in intracellular parts of the protein^8^. Though the frequencies vary for different ethnicities, they are common in the general population; In fact, the alternative allele is present in 57%, 55% and 49% of genotypes for T1236C, T2677G and T3435C respectively^11^. The three also make a common haplotype, occurring in 33% of individuals^12^.

Variants, particularly ones defined as “silent” such as synonymous alterations, can modify the gene expression process. They can affect transcription through changing transcription-factor binding sites (TFBS)^13^; they can impact translation and co-translational folding through changing codon usage^14,15^; they can also accelerate mRNA degradation by changing affinity to miRNA binding sites^16^ or lead to mis-spliced proteins by modifying the nucleotides in the vicinity of donor or acceptor sites^17^. Altogether, they can impact all phases of gene expression. Though these three variants are widely studied, conclusions regarding their effects have been exceptionally inconsistent^18^. Some studies suggest they change mRNA or protein expression^19–23^ while others claim they do not^24–29^ and associate their presence with modifications in protein structure^24,29^. Some find them to significantly impact patients’ response to chemotherapy and survivability while others do not^30–46^. Remarkably, controversy is also found among those who claim that the variants do affect survivability^47^; whether the variants are detrimental or advantageous for patient survival remains a matter of disagreement.

The effects of these MDR1 variants on p-gp expression, function and on chemotherapy resistance have been studied mostly in clinical trials and in-vitro cell line experiments. Each method has inherent disadvantages; results of clinical trials are affected by factors such as cohort size, patient ethnicity, tumor type and conducted chemotherapy regimen. In-vitro experiments cannot fully mimic the symbiosis of a cell with its natural environment and substantially differ from the corresponding cell type in-vivo^48^. Even though the effects of the variants may vary under different conditions, it is possible that some of the contradictions emerge not due to biological complexity but rather due to impediments in scientific methods. The objective of this paper is to examine, for the first time, the effects of T1236C, T2677G and T3435C on all phases of the gene expression process using various computational approaches. We aim to gain a better understanding of the mechanism of action of this three MDR1 variants, endeavoring to propose plausible explanations and validate some of the previously proposed effects.

## Methods

### Data sources

For performing the various analyses, we utilized data of several known databases:

#### TCGA

The Cancer Genome Atlas holds genetic, epigenetic, clinical and gene expression data of tens of thousands of cancer patients. The data was downloaded from TCGA (https://www.cancer.gov/tcga) on November 2021. Specifically, data of simple nucleotide variation (SNVs – small nucleotide variations), of MDR1 mRNA expression and clinical records regarding patients’ vital status were used for this study.

Note that the patients’ genotypes are confidential. Therefore, we know whether a patient acquired a mutation or not but we do not know the patient’s second allele. For example, we know whether a patient is T1236C positive or not but the second unmutated allele could contain either a “C” or a “T”.

#### ENSEMBL

Ensembl^49^ provides a centralized platform for the storage, analysis, and visualization of genomic data. For CAI and MFE measurements, the complete human CDS was downloaded from ENSEMBL (https://ftp.ensembl.org/pub/release-109/fasta/homo_sapiens/cds/) on 2020.

#### PaxDb

PaxDb^50^ is a protein abundance database that contains whole genome protein abundance information across organisms and tissues. For the CAI calculation the protein levels of human (whole organism and from integrated experiments) were downloaded from the PaxDb website on 2020 (https://pax-db.org/dataset/9606/1502934799/).

### MDR1 expression

#### TCGA

To examine the effect of T1236C, T2677C, T3435C and all their possible combinations on MDR1 levels of cancer patients we used TCGA SNVs and expression data. At the time of data curation there were 15,500 patients with both available SNV information and MDR1 mRNA expression levels. For each variant or haplotype we performed the following –

- Split the patients to two categories: those who acquired the variant/haplotype in their cancerous tissue sample and to those who did not. The first group (carriers) contains several dozens of individuals or less (depending on the variant) and the latter (non-carriers) contains more than 10,000 individuals.

- Randomly chose a sub-group from the non-carriers group. The size of the sub-group is the size of the carriers group.

- Repeated the random sampling for 100,000 times to get 100,000 sub-groups of non-carriers.

- For each of the non-carriers’ sub-groups and the carriers’ group, obtained the mean MDR1 expression (in FPKM) of the group.

- Obtained an empirical p-value by comparing the mean MDR1 expression of the carriers group to the distribution of mean MDR1 expression on the non-carriers groups.

To assert that the results are not caused by differences in tumor mutational burden (TMB), we repeated the analysis when controlling for this confounder; the balance of the TMB between the mutated and control groups was tested using a two sample KS test^51^. While T1236C mutated and control groups were balanced, restriction was used to balance The T2677G and T3435C mutated and control groups.

Controlling for TMB did not change the outcomes of the analysis.

### Enformer

To examine the effect of T1236C, T2677C, T3435C on MDR1 levels we used Enformer^52^. The input of the model is a sequence of length 393,216 nucleotides and the variant is positioned in the middle (nucleotide 196,608). The centered 114,688 nucleotides are in the prediction range and the rest are provided as context. The output of the model is a matrix with 5,313 rows that represent experiment tracks and 896 columns that represent bins. Each track in the output matrix is the prediction of gene expression under specific conditions (for example, CAGE measurements from liver tissue or DNASE measurements from the brain tissue). Each bin represents 128 nucleotides, allowing the 896 bins to represent expression for the full prediction range. For each variant we used Enformer four times; We input the reference and mutated sequence for both the forward and reverse strands, all centered around the position of the variant. Multiple approaches were taken to derive a single score from the Enformer output, such that the score will capture the difference in MDR1 expression that is caused by the variant. The following steps were performed:

- The absolute difference in expression was calculated for each strand using the following formula:

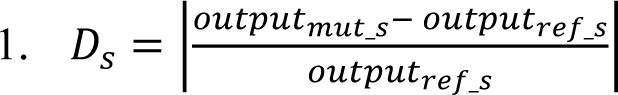

Where 𝑜𝑢𝑡𝑝𝑢𝑡_𝑚𝑢𝑡_𝑠_ is the Enformer output matrix given the mutated sequence of strand 𝑠, 𝑜𝑢𝑡𝑝𝑢𝑡_𝑟𝑒𝑓_𝑠_ is the Enformer output matrix given the reference sequence of strand 𝑠 and 𝐷_𝑠_ is the matrix of absolute differences of strand 𝐷_𝑠_. The output of this step is comprised of two matrices of size (5313, 896), one for each strand.

- An average across all bins was performed. The output of this step is comprised of two matrices of size (5313, 1).

- Merging across tracks was performed using several methods –

- An average across all tracks
- An average across all CAGE tracks
- An average across tracks associated with tissues where MDR1 is highly expressed
- An average across CAGE tracks associated with tissues where MDR1 is highly expressed The output of this step (for each method separately) is comprised of two scores, one for each strand.

- An average across both strands was performed. The output of this step is the final score representing the change in expression caused by the variant.

The same process was performed for random variants with similar characteristics (explained in the “Empirical p-values” section) and the scores of the original and random variants were compared. If the score of the variant was larger than the scores of 95% of the random variants it was deemed to significantly change MDR1 expression.

### Splicing events

To examine the effect of T1236C, T2677C, T3435C on the occurrence of splicing events we used SpliceAI^53^. The input of the model is a sequence the length of 11,001 nucleotides; the position of the variant is in the middle of the sequence, the proximal 500 nucleotides from each side are in the range of the prediction and the distal 5000 nucleotides from each side are provided as context. For each nucleotide that is in the prediction range, the model predicts its probability of being a donor site and its probability of being an acceptor site. We perform the prediction for both the original sequence and mutated sequence, centered around each of the three variants. For each position in the prediction range, we calculated the difference in the probability of it being a donor/acceptor site that was caused by the variant. We searched for positions for which the probability of being a donor or acceptor site changed by more than 50% . Positions exhibiting an increment of more than 50% are considered new prospective donor/acceptor sites, whereas positions for which the probability decreases by more than 50% are regarded as potentially abolished donor/acceptor sites.

### Translation rates

To examine the effect of T1236C, T2677C and T3435C on translation rates we utilized several measures positively correlated with it - MFE, CAI, FPTC and tAI. We calculated these measures for both the original and mutated MDR1 CDS sequences and examined the difference in these measures at the position of the variant.

#### MFE

A per-position MFE score was computed using ViennaRNA^54^; first, a sliding window (length = 39 nucleotides, stride = 1 nucleotide) was used to obtain a per-window MFE score. Then, the MFE score of a specific position in the CDS was set as the average of all MFE scores of the windows that the position is in.

#### CAI

Human CAI weights were computed as suggested in the original paper^55^. The set of highly expressed genes (15% most highly expressed) was curated using human protein expression levels from PAXdb^50^.

#### FPTC

Human FPTC weights were downloaded from the Kazusa website^56^.

#### tAI

tAI tissue-specific weights were taken from Hernandez-Alias et al.^57^ and the s weights were optimized as depicted in Sabi et al.^58^

### Co-translational folding

To examine the effect of T1236C, T2677C and T3435C on the co-translational folding of p-gp, a computational model that assesses which positions in the CDS are important for correct protein folding was deployed. The model is yet to be published and therefore we will provide much detail about the model’s methodology. The analysis is based on the basic assumption that co-translational folding is governed by local translation rates, and that this rate would be evolutionary conserved for a position that is crucial for correct folding^59^ . Thus, it searches for positions with both evolutionarily conserved low and evolutionarily conserved high MFE (a measure correlated with translation rate) across orthologous versions of a gene. For a given gene (MDR1 for example) the conserved positions are found in the following manner-

- The human CDS sequence, along with all available orthologous CDS sequences are downloaded from Ensembl (https://rest.ensembl.org/documentation/info/homology_ensemblgene). Both the nucleotide and respective amino-acid sequences are downloaded for each ortholog. Specifically, for MDR1 - 383 orthologs were found.

- Using the nucleotide sequences, an MFE score is calculated for each position in each orthologous sequence as was depicted in the “Translation rates” section.

- To find conserved positions we must first align them; the orthologues amino-acid sequences are aligned using Clustal Omega^60^ to create a multiple sequence alignment (MSA) for the gene.

- According to the amino-acid alignment, the nucleotide sequences are also aligned with Pal2NAl^61^.

- The MFE scores that were obtained for each position in each nucleotide sequence are now mapped to their appropriate location in the nucleotide alignment.

- An average MFE score across orthologs is calculated per CDS position. Currently, we get a single MFE score for each position in the CDS, indicating the average MFE at this position across the different organisms.

- Computing significance- to find the MFE scores that are significantly lower or higher than expected by chance we create permuted versions of the MSA, calculate the average MFE per position for the permuted versions and compare them to the scores of the original version.

- In the first type of permuted version of the MSA, called “vertical permutation”, we shuffle synonymous codons within the same MSA column. In the second type of permuted version of the MSA, called “horizontal permutation”, we horizontally swap between synonymous codons of pairs of columns. Both methods affect the MFE scores while keeping basic characteristics of the MSA such as the amino-acid, codon and nucleotide content. One hundred permutations of the MSA are created for each kind.
- We calculate the per-position MFE score averaged across orthologs for each permuted MSA in the same manner as was done for the original MSA. At this point, for each CDS position we have a single true MFE score and 100 scores from each of the permuted versions.
- For each position we obtain a z-score and respective p-value for each permutation type.
- We perform an FDR correction for the p-values.
- Finally, we intersect the results to get positions that had significantly low MFE and positions that had significantly high MFE when compared to both kinds of permuted versions of the MSA. We require both the vertical and horizontal p values to be equal or smaller than 0.05.

### Survivability of TCGA patients

To examine the effect of T1236C, T2677C, T3435C and all their possible combinations on the survival of cancer patients we used TCGA SNV and clinical data. At the time of data curation there were 11,086 patients with relevant clinical information. We used the vital status of the patients and a matching time-stamp in order to create Kaplan-Meier survival curves^62^ for the carriers and non-carriers groups. The time-stamp was derived from the maximum value of the following attributes (not all attributes were available for each patient)-

- Days from the initial diagnosis to current follow-up
- Days from the initial diagnosis to the current confirmation of vital status
- Days from the initial diagnosis to patient death

The logrank test^63^ was used to assess whether the survival curves of the carriers and non-carriers group significantly differ.

As in the MDR1 expression analysis, balancing the TMB did not affect the outcome for either of the three variants.

### Empirical p-values

To infer the significance of the changes caused by T1236C, T2677C and T3435C in several of the analyses, they were compared to changes caused by random variants with similar characteristics. T2677G is a non-synonymous variant and therefore its effect was compared to other, randomly sampled, T->G non-synonymous variants in the MDR1 CDS. T1236C and T3435C are synonymous variants and therefore were both compared to randomly sampled synonymous T->C variants in the MDR1 CDS. Each original variant was compared to 100 randomized sequences.

### TCGA variants correlated with T1236C

We investigated the associations between the occurrence of somatic variants T1236C, T2677G, and T3435C in the genomes of TCGA cancer patients, with regards to both MDR1 expression and survival outcomes. Significant correlations were found for T1236C. To investigate the possibility of a causal relationship, we further examined the influence of other variants highly correlated with T1236C on MDR1 expression and patient survival, aiming to eliminate their potential effects. Highly correlated variants were defined as variants that are detected in the genomes of more than 75% of T1236C positive patients.

## Results

Diverse computational models were deployed aiming to assess whether T1236C, T2677G and T3435C could modify different phases of the gene expression process (figure S1). Additionally, genetic, clinical and expression data from The Cancer Genome Atlas (TCGA) were analyzed for this purpose. It is important to mind the difference between a germline variant, a genetic variation that is inherited and may be prevalent in the population, and a somatic variant which is a genetic variation that is acquired during one’s lifetime. Note that when analyzing TCGA data we evaluate the effect of somatic variants in cancerous tissues, whereas the computational models can be used to interpret the effects of both somatic and germline variants.

### T1236C potentially increases mRNA levels via an undetected mechanism

MDR1 abundance was compared between TCGA patients that acquired any of the three somatic variants and patients who did not (see Methods). Results, shown in figure 1, indicate marginal association between T1236C and MDR1 expression, with a two-fold increase (3.26±8.4 FPKM for the non-carriers and 6.54±12.98 FPKM for the carriers, p = 0.07). This suggests that the presence of a “C” allele in position 1236 is correlated with increased expression of MDR1. When examining the effects of other TCGA variants that are highly correlated with the presence of T1236C (see Methods) on MDR1 expression, the increase is much smaller (fold change ranges between 1.01 to 1.05. See figure S2), implying that these variants are not the cause for the detected two-fold increase. Additionally, all the variants that are highly correlated with T1236C were found to be in MUC6, a gene that is prevalently mutated in cancer patients and is not known to be related to MDR1 regulation. Differences in mRNA abundance between carriers of the haplotypes and non-carriers were found non-significant (0.11 < p < 0.24) (figure S3). However, this could be due to the loss of statistical power (20 patients carry the T1236C variants whereas only five carry all three variants).

**Figure 1:**
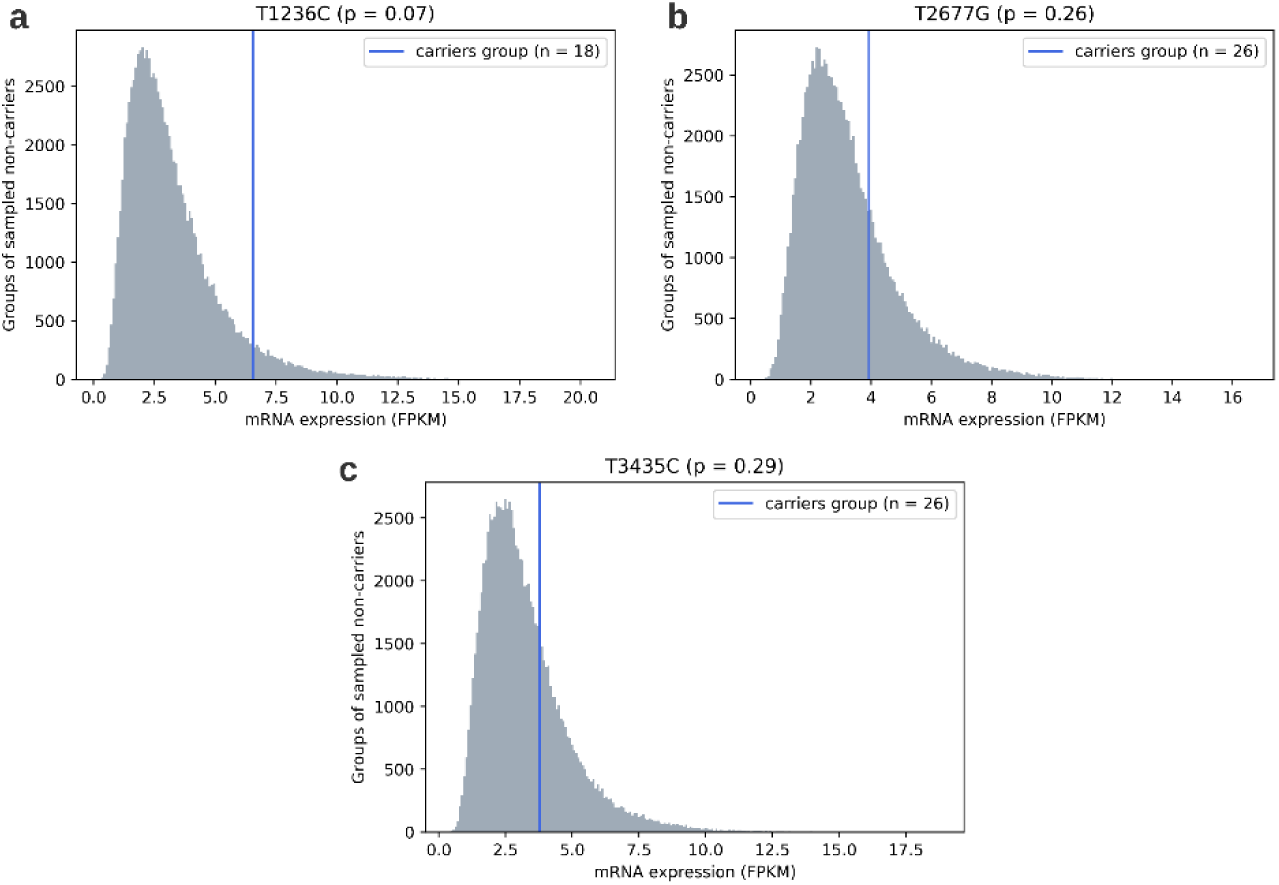
MDR1 expression levels of carriers vs. non-carriers of the three variants in the TCGA database. a: T1236C ; b: T2677G ; c: T3435C .Comparison of the mean MDR1 expression of the carriers group (vertical line) to the mean MDR1 expression levels of 100,000 groups of randomly chosen non-carriers (distribution). Size of the carriers’ group and non-carriers’ groups are the same.

T1236C, T2677G and T3435C are far from the transcription start site (TSS) with a distance of 50.6, 59.5 and 91.5 kilobases respectively. Additionally, they are not located in known enhancers. Therefore, they are not expected to impact transcription. If T1236C does cause an increase in MDR1 expression it is more likely due to modifications in mRNA stability and degradation.

We used Enformer^52^ to computationally assess whether the variants are likely to cause a change in MDR1 expression. Enformer is a transformer-based neural network that is trained on thousands of epigenetic and transcriptional datasets to predict gene expression. It identifies complicated genomic patterns related to gene expression regulation such as TSSs, TFBSs, histone modifications sites and miRNA binding sites.

With an input sequence of 393,216 nucleotides, it is currently the gene expression predictor with the widest receptive field. For each variant we ran Enformer four times – on the reference sequence and on the mutated sequence, for both strands (see Methods). The predicted change in MDR1 expression was not significant for any of the variants, including T1236C. It is possible that T1236C does not cause the detected increase in MDR1 expression but is simply correlated with it. Another possibility is that T1236C does affect MDR1 expression through a mechanism that is not detected by Enformer (e.g. mRNA stability). Perhaps, due to the variant’s large distance from the TSS^64,65^.

### T1236C, T2677G and T3435C do not seem to affect mRNA splicing

SpliceAI^53^ was used to assess whether the variants change MDR1 splicing. Given a sequence, the output of spliceAI is a matrix of probabilities, indicating the probability of each position in the sequence to be a donor or acceptor site (see Methods). For each variant we ran SpliceAI twice – on the reference sequence and on the mutated sequence. Using the difference between probabilities we searched for canceled or newly generated donor and acceptors sites, as these events could lead to alternative splicing.

None of the variants were found to cause significant changes.

### T1236C, T2677G and T3435C potentially increase p-gp levels through raising global translation rates

The rate of translation is a key component that shapes protein abundance. A change in the translation rate of a single codon can impact protein abundance if it is located in a translation bottleneck. The translation rate varies throughout the mRNA sequence and is dependent on multiple cellular conditions such as accessibility of the mRNA to the ribosome, codon usage bias and availability of tRNAs. In this section we examine several measures correlated with the translation rate and evaluate whether the three variants are expected to change it, locally or globally.

### T1236C, T2677G and T3435C all decrease mRNA folding strength. T1236C and T3435C potentially increase global translation rates due to this decrease

Minimum Free Energy (MFE) is a measure derived from predictions of the secondary structure of an mRNA sequence; a lower MFE value indicates a structure that is tightly packed and less accessible for translation. Therefore, lower MFE values are correlated with lower translation rates^66^. We computed MFE scores for the region surrounding each of the three variants (see Methods) and predicted the instigated change in MFE. The results (figure 2) demonstrate that all three variants cause an increase in MFE. This increase is significantly larger (p = 0.01, p = 0.02 and p = 0.04 for T1236C, T2677G and T3435C respectively) than the increase caused by random variants with similar characteristics in the MDR1 gene (see Methods). Moreover, positions 1236 and 3435 have relatively low MFE scores compared to all coding sequence (CDS) positions (19^th^ and 12^th^ percentile respectively) and therefore make constitute possible translation bottlenecks (figure S4), suggesting that T1236C and T3435C could increase global translation rates.

**Figure 2:**
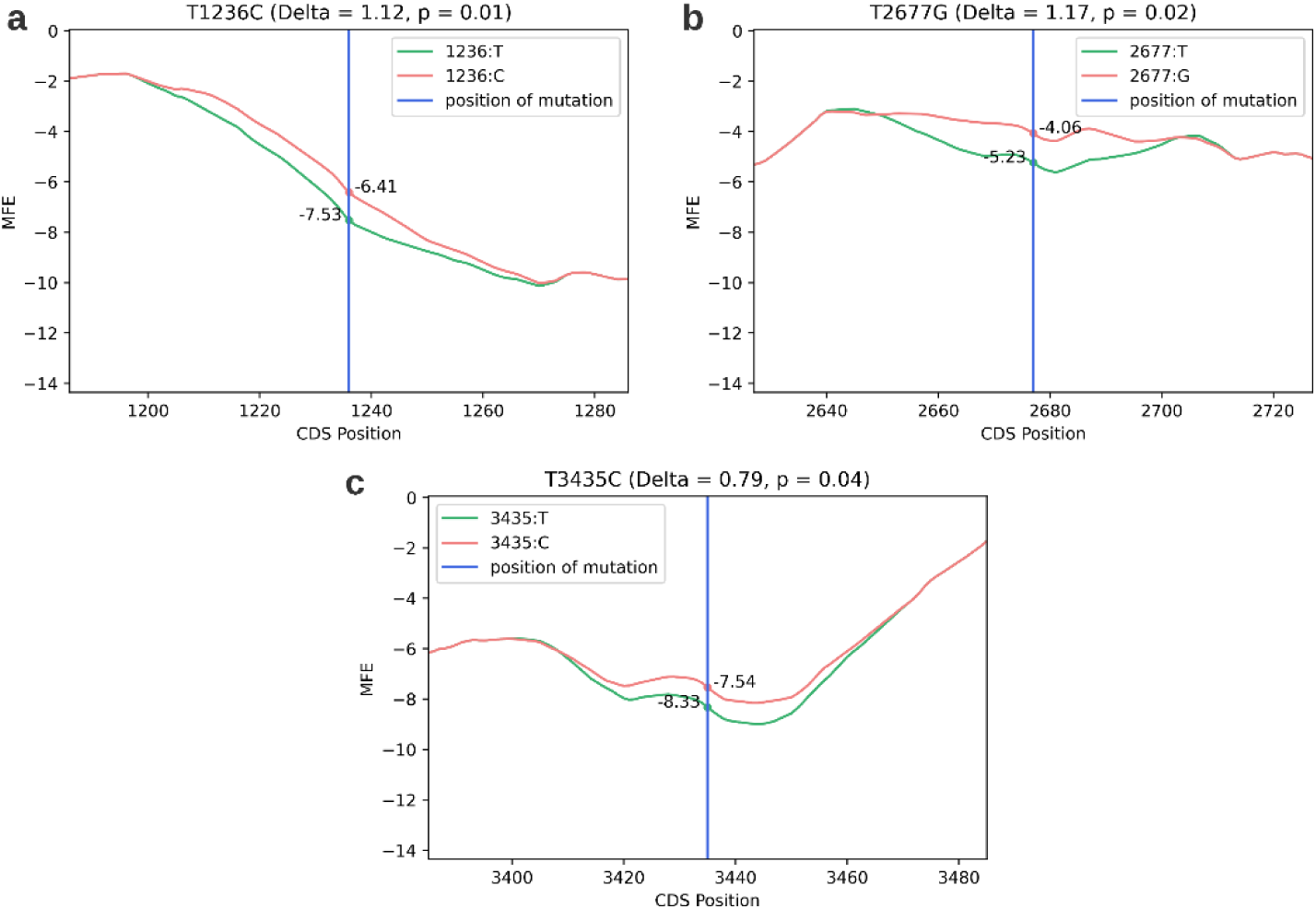
Effect of the three variants on MFE in their vicinity. a: T1236C ; b: T2677G ; c: T3435C . x axis: the nucleotide position in the coding sequence (CDS); y axis: MFE score. The vertical line indicates the position of the variant, and the curves depict the MFE scores of the nucleotides proximal to the position of the variant, with (pink) or without it (green).

### T1236C, T2677G and T3435C increase the optimality of codon usage. T1236C potentially increases global translation rates due to this increase

We compute Codon Adaptation Index (CAI) and Frequency Per 1000 codons (FPTC) to measure the change in codon usage bias (CUB) caused by the three variants, where CAI is computed for the synonymous variants and FPTC for the non-synonymous variant. Higher CUB suggests better adaptation of the sequence to the cellular translation machinery and resources and is correlated with faster translation^67^. Results (figure 3) indicate that all three variants cause a less prevalent codon to be replaced by a more prevalent codon, suggesting an increase in local translation rate. Moreover, T1236C substitutes the least prevalent codon of Glycine to the most common one. When comparing to random variants with similar characteristics in the MDR1 gene, the increase in CAI caused by T1236C is marginally significant (p = 0.08). Also, the CAI score of codon 412 (in which position 1236 resides) is in the 16^th^ percentile of CAI scores compared to all the codons in the CDS, further strengthening the possibility that it is a translational bottleneck and that T1236C could increase global translation rate.

**Figure 3:**
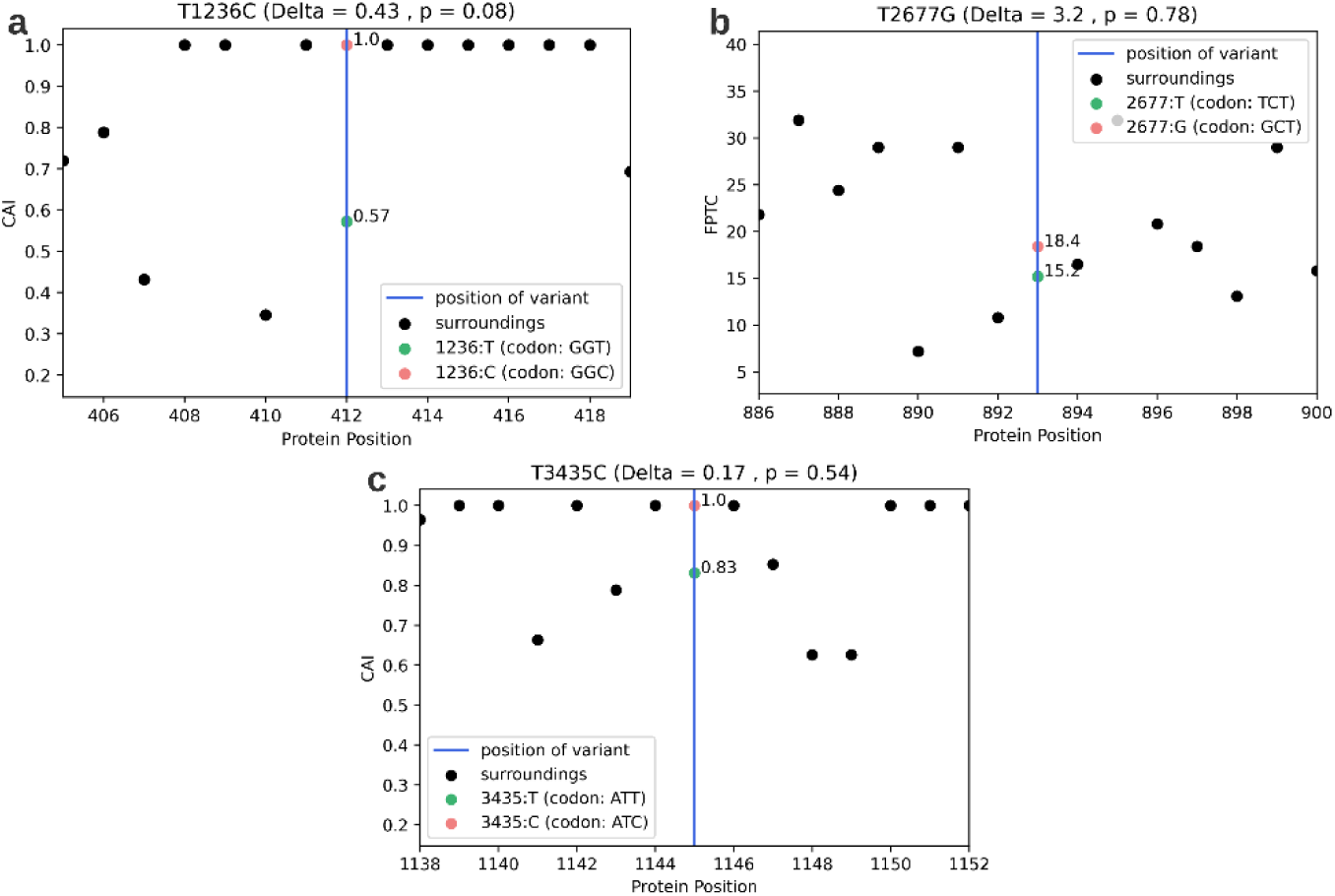
Effect of the three variants on CUB. a: T1236C ; b: T2677G ; c: T3435C . x axis: the amino-acid position in the protein sequence; y axis: the CUB score (CAI/FPTC). The vertical line indicates the position of the variant. Black dots indicate the CUB scores in the vicinity of the variant and the dots on the vertical line indicate the CUB score of the mutated position, before (green) and after (pink).

### T1236C and T2677G improve adaptation to the tRNA pool in tissues where MDR1 is expressed. T2677G potentially increases global translation rates due to the improvement in adaptation

The tRNA Adaptation Index (tAI) is a measure of translational efficiency which considers the intracellular concentration of tRNA molecules and the efficiencies of each codon–anticodon pairing^68^. Higher tAI scores are given to codons with better tRNA availability and are correlated with a higher translation rate^69^. The tRNA availability changes substantially between different tissues and organs. We examine the effect of the variants on the tAI profile in tissues where the MDR1 gene is typically expressed (see Methods) – liver, kidney, colon and brain tissues. Figure 4 demonstrates the effect of the variants on the tAI profile in the liver tissue, but it is similar for all other examined tissues (figures S5-S7). The results of this analysis show that both T1236C and T2677G cause an increase in tAI. Moreover, T2677G replaces a codon with an extremely low tRNA availability to a codon with much higher tRNA availability, in all examined tissues. When comparing the tAI change caused by T2677G to changes caused by random variants with similar characteristics in the MDR1 gene, the former is found either significantly or marginally significantly larger in all relevant tissues (p <= 0.1). Moreover, the tAI score of the codon affected by T2677G is in the 1^st^-5^th^ (depending on the tissue) percentile of the tAI scores of all the codons in the CDS, strengthening the indication that the position could be a translation bottleneck and that T2667G could increase global translation speed.

**Figure 4:**
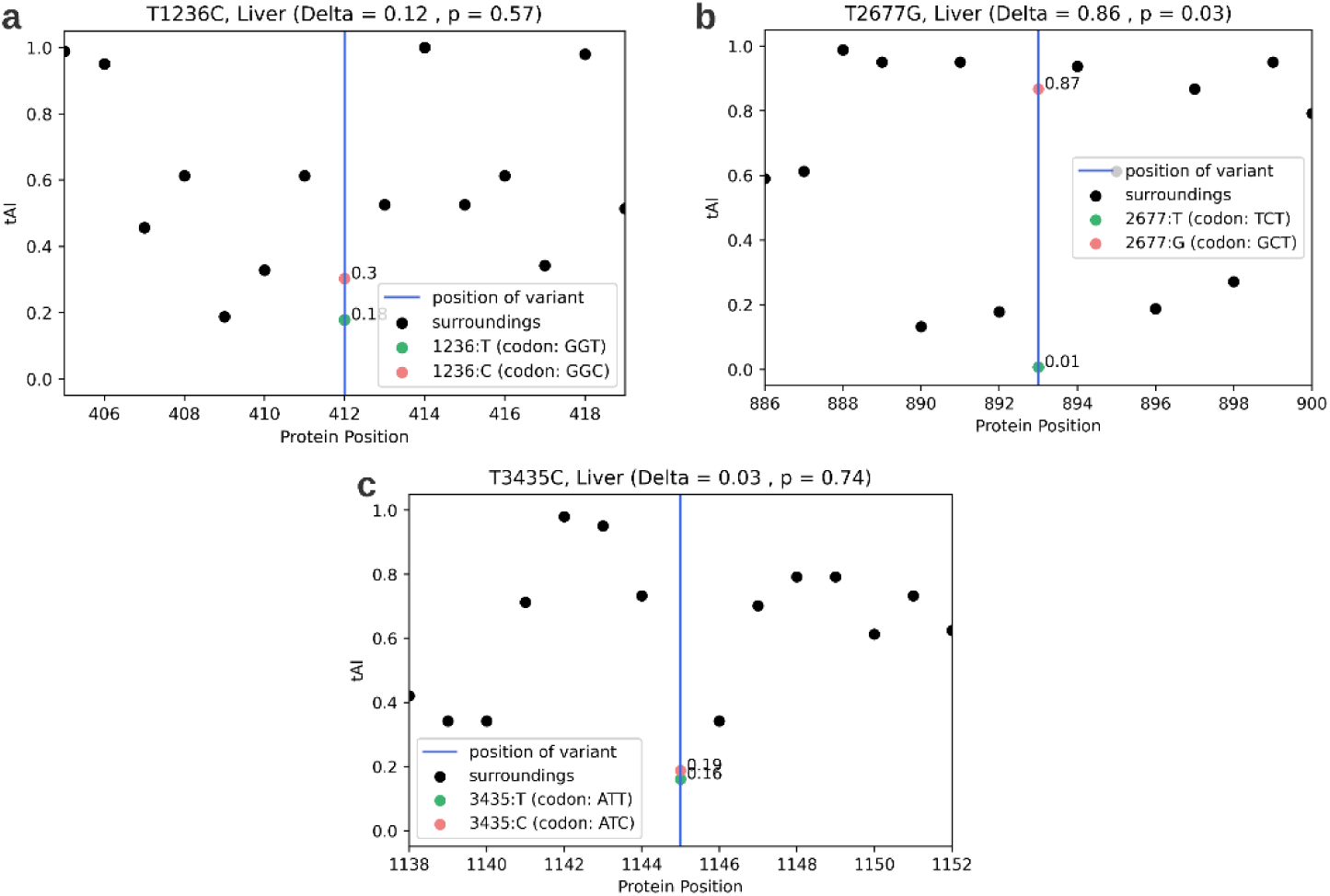
Effect of the three variants on tAI. a: T1236C ; b: T2677G ; c: T3435C .x axis: the amino-acid position in the protein sequence; y axis: the tAI score. The vertical line indicates the position of the variant. Black dots indicate the tAI scores in the vicinity of the variant and the dots on the vertical line indicate the tAI score of the mutated position, before (green) and after (pink).

To conclude, the variants are in positions that are potential translation bottlenecks as they all obtain scores that are close to the minimal score of the entire CDS in at least one of the examined measures. All three variants lead to an increase of MFE and CUB scores, the increase in MFE being significantly larger than expected by chance. Additionally, T1236C and T2677G cause an increase in tAI scores in tissues where MDR1 is expressed, with T2677G leading to an extreme and significant tAI change. These results suggest that the variants likely increase local translation rates in their vicinity, and possibly increase global translation rates and protein levels.

### T3435C potentially modifies co-translational protein folding through raising local translation rate in a conserved slowly translated region

Co-translational folding is the mechanism in which the nascent protein begins to fold during its translation^70^. This process was shown to be governed by local translation rates^59^. To examine whether T1236C, T2677G and T3435C influence co-translational folding we deployed a model which detects positions that have evolutionary conserved extreme MFE scores. The rationale being that MFE scores can be used as proxies for translation rates and that positions with evolutionary conserved extreme translation rates (especially slow rates) are largely conserved as such due to their importance for optimal co-translational folding. Therefore, it is possible that variants in these positions interfere with the co-translational folding mechanism. In order to find positions in the CDS of MDR1 with conserved extreme MFE scores, the model utilizes orthologous MDR1 genes from hundreds of organisms, calculates their MFE profiles and compares them to the MFE profiles of permuted versions of these genes (see Methods). Model output (figure 5) indicates that T3435C is in a position with evolutionary conserved low MFE, surrounded by a stretch of positions with conserved low MFE (6 nucleotides upstream of T3435C and 17 downstream of it). Combining the results of this model and the results of the previous analysis which suggests that T3435C causes a local increase in translation rate, we deduce that it is possible that T3435C causes an increase in translation rate in a region of conserved low translation rates, and thus modifies co-translational folding. Both T1236C and T2677G were not found to be in a position of evolutionary conserved extreme MFE.

**Figure 5:**
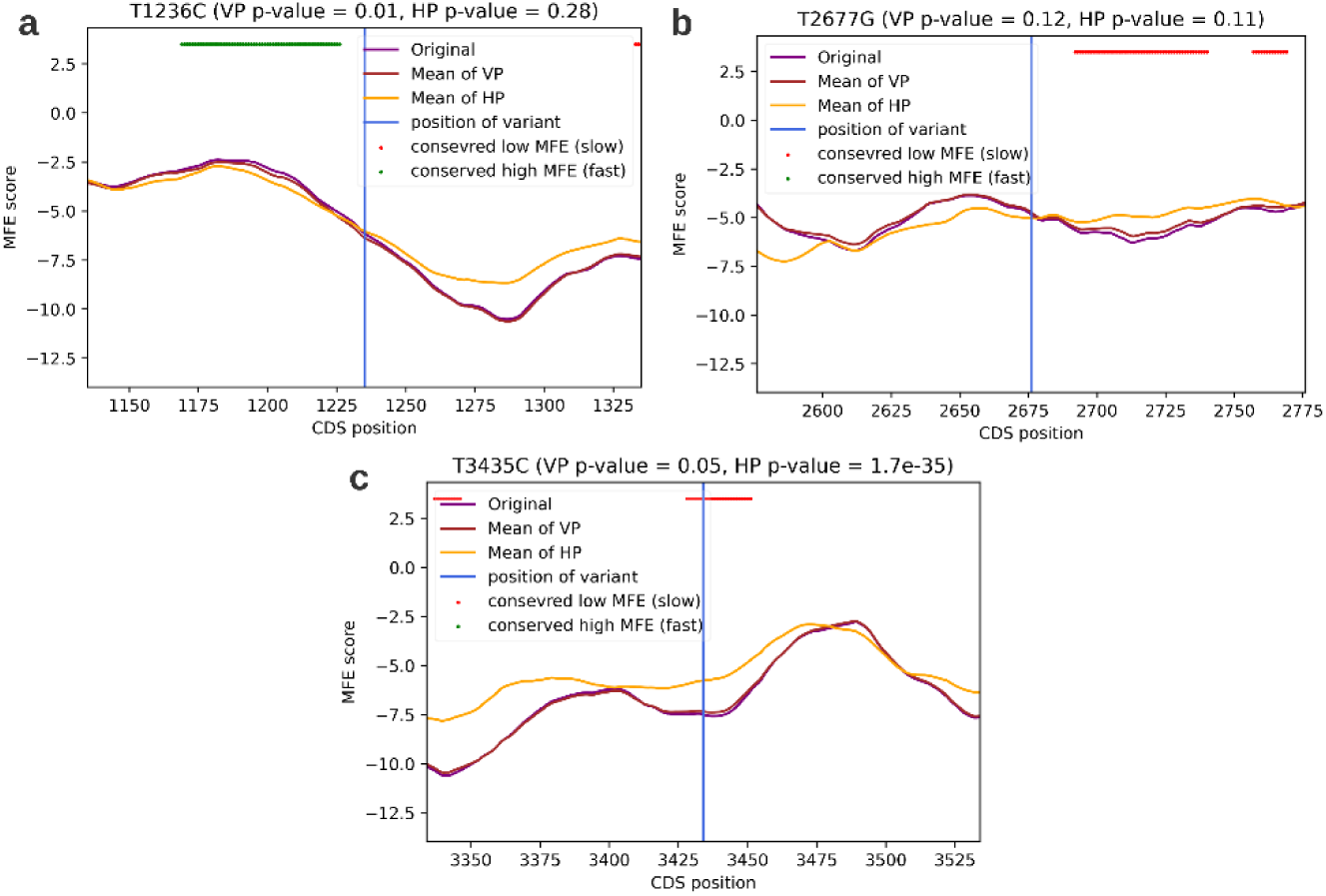
Regions of evolutionary conserved low or high MFE in the vicinity of the three variants. a: T1236C ; b: T2677G ; c: T3435C . x axis: the nucleotide position on the CDS of the MDR1 gene. y axis: the MFE score, which is used by this model as the proxy for translation rate. The purple curve indicates the mean MFE score of the true MDR1 CDS across 383 orthologs. The orange and brown curves indicate the mean MFE score of a permuted MDR1 CDS, using two different kinds of permutations – vertical permutations (VP) and horizontal permutations (HP). Green dots are nucleotide positions for which the MFE of the true MDR1 was significantly higher than the MFE of both permutated versions, across organisms. Red dots are nucleotide positions for which the MFE of the true MDR1 was significantly lower than the MFE of both randomized versions, across organisms.

### T1236C potentially decreases patient survivability

Clinical data of TCGA patients were used to assess the variants’ effect on survivability. Kaplan Meier curves^62^ were compared between patients that acquired one or more of the three variants in their cancerous tissue and those who did not (see Methods). Results (figure 6, figure S8) suggest that T1236C decreases survival probability (p = 0.02) whereas T2677G and T3435C do not. Other TCGA variants that are highly correlated with the presence of T1236C (see Methods) are not correlated with decreased survivability (figure S9), enhancing the likelihood that T1236C is the causal variant.

**Figure 6:**
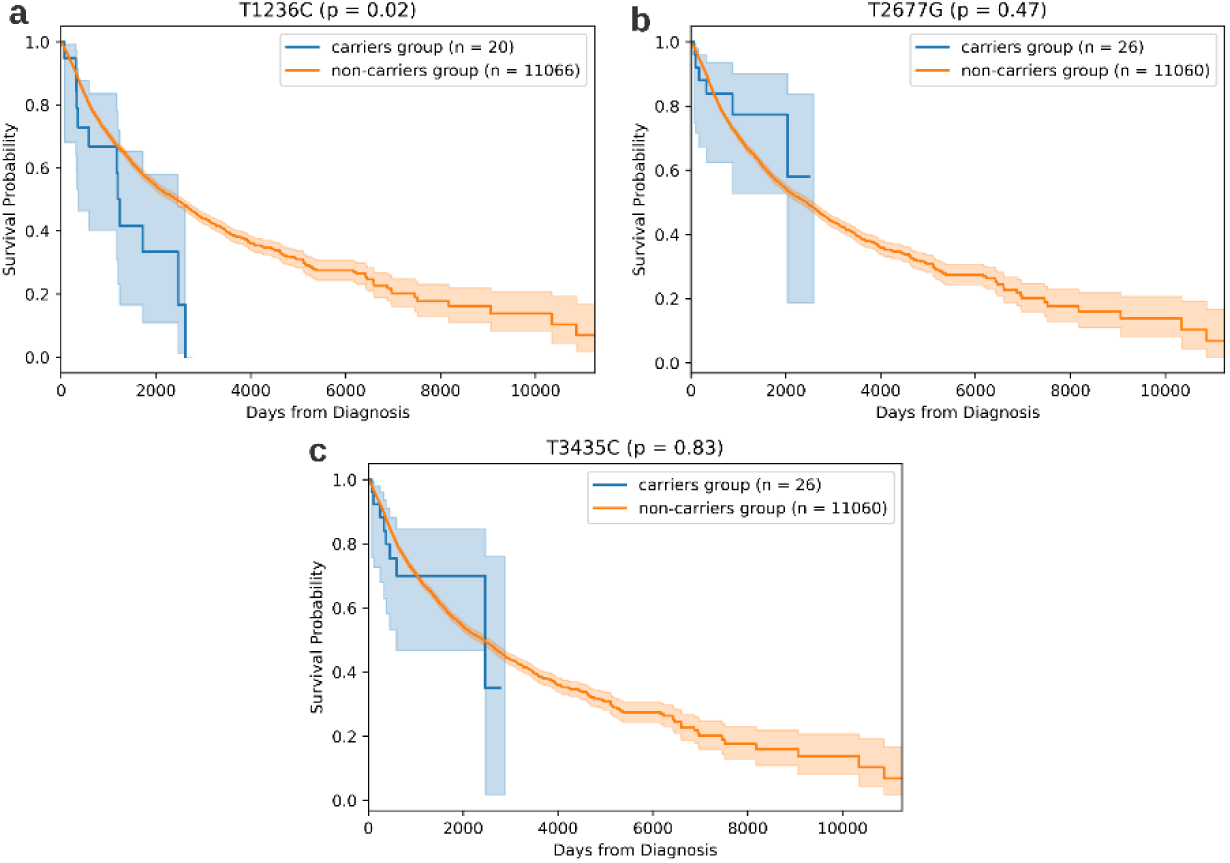
Effect of the three variants on patient survivability. a: T1236C; b: T2677G; c: T3435C. Comparison of the Kaplan-Meier survival curves of the carriers’ group and the non-carriers group of the three variants.

Due to p-gp’s role in chemotherapy resistance, we hypothesized that the effect of T1236C on survivability would be even more significant in patients that have undergone chemotherapy treatments, but this signal was not detected (figure S10). However, it is possible that the signal is not detected due to a reduction of statistical power. To conclude, the computational results support a hypothesis that T1236C could decrease patient survivability.

## Discussion

Since the discovery of MDR1 and its many variants, numerous in-vitro and in-vivo studies have attempted to understand their mechanisms of action and contribution to chemotherapy resistance and cancer prognosis. Due to contradictory findings and the lack of success of MDR1 inhibitors in clinical trials^71^, research of MDR1 and its variants has gradually diminished. However, the current surge in genomic data and evolving technology enables the examination of this case from a new, data-driven, perspective.

Our analyses aid to paint a clearer image, providing evidence of multiple accumulating effects. T1236C seems to significantly increase MDR1 expression. Additionally, it causes an increase in CAI, tAI and MFE, all positively correlated with translation rates. Because it is in a region of low MFE and CAI, this increase could lead to a rise in global translation rates and p-gp over-expression. Moreover, T1236C was found to be correlative with worse patient prognosis, which can be explained by overexpression of the protein. T2677G Also causes an increase in measures correlated in translation rates and most significantly effects tAI. It causes a codon that is extremely rare in tissues where MDR1 is expressed to be replaced with a common codon. T3435C also increases MFE and CAI in a region of very low MFE. Additionally, it possibly modifies p-gp structure through changing local translation rate. Altogether, when exploring the mechanism of action of T1236C, T2677G and T3435C, it is much more likely that they cause an increase in p-gp expression rather than a decrease, providing an advantage for the cancer cell.

Though the findings of previous research on the matter are inconsistent, our analyses support the conclusions of several major studies. Kimchey-Zarfaty at el. demonstrated that T3435C, combined with either T1236C or T2677C, changes MDR1 substrate specificity^24^. They hypothesized that T3435C modifies co-translational folding (and therefore protein conformation) because it changes the local translation rate in a slowly translated region, important for correct co-translational folding. They supported their hypothesis by identifying a cluster of non-prevalent codons in the vicinity of T3435C (as seen in sub-figure 3c). Our model, which utilizes MFE and assesses which positions are evolutionarily conserved for slow or fast translation, indeed detects 3435 in a region that is likely conserved for slow translation, supporting their findings. Moreover, we demonstrated that T3435C increases both MFE and CAI scores, suggesting that it could increase the local translation rate in this slowly translated region and thus modify co-translational folding. Kimchey-Zarfaty et al. also examined mRNA expression, protein expression and aberrant splicing events in wild-type and polymorphic p-gp and report no aberrant splicing events. To the best of our knowledge, no prior study has suggested that T1236C, T2677G or T3435C cause alternative splicing. Our results, obtained using SpliceAI^53^, support the consensus. As for mRNA and protein expression, Kimchey-Zarfaty et al. report equivalent levels for wild type and polymorphic variants. In fact, most in-vitro studies do not report a significant change, while some in-vivo studies do.

This could be due to a complicated mechanism of action, that cannot be detected out of the natural cellular environment. Nevertheless, reports of the in-vivo studies are also controversial, perhaps due to the tremendous variability in MDR1 expression between patients, small cohorts and difference in patient ethnicities, genetic backgrounds and tumor types. Combining the results of our analyses, we assess that if the variants do modify mRNA and protein levels, they should cause an increase rather than a decrease. In another comprehensive study, Johnatty SE et al.^72^ examine 4,616 ovarian cancer patients from the Ovarian Cancer Association Consortium (OVAC) and TCGA, that have received chemotherapy treatments. They found a marginal association of T1236C with worse overall survival and did not find association between T2677G or T3435C and survival parameters. These findings are supported by our pan-cancer survival analysis on TCGA data, shown in figure 6.

When reviewing the analyses described in this study, we must remember the limitations of in-silico models as well; It is challenging to create models that capture all relevant processes and interactions in highly complex biological mechanisms such as gene expression regulation. Additionally, some of the models were trained on data obtained from wet lab experiments and thus are also subjected to noise.

Nonetheless, computational models have many advantages; they are often less expensive, faster and can incorporate larger quantities of data than in-vitro experiments. We believe the incorporation of computational, data-driven models is both essential and highly beneficial when analyzing the complex effects of variants on gene expression regulation, generally and specifically for the case of MDR1.

## Data Availability

The code is available at https://github.com/Talgutman/MDR1_variants

## Supplementary Data Statement

Supplementary Data are available at NAR online.

## Data Availability

All data produced in the present study are available upon reasonable request to the authors

## Acknowledgements

We thank Alma Davidson, Yoram Zarai, Sanjit Batra and Yun S. Song for their contributions in different analyses of this study.

## Bibliography

1. Juliano, R. L. & Ling, V. A surface glycoprotein modulating drug permeability in Chinese hamster ovary cell mutants. BBA - Biomembranes 455, 152–162 (1976).

2. Allikmets, R., Gerrard, B., Hutchinson, A. & Dean, M. Characterization of the human ABC superfamily: isolation and mapping of 21 new genes using the expressed sequence tags database. Hum Mol Genet 5, 1649–1655 (1996).

3. Li1, Y., et al. The structure and functions of P-Glycoprotein. Curr Med Chem 17, 786–800 (2010).

4. Sakaeda, T., Nakamura, T. & Okumura, K. MDR1 genotype-related pharmacokinetics and pharmacodynamics. Biol Pharm Bull 25, 1391–1400 (2002).

5. Thiebaut, F. et al. Cellular localization of the multidrug-resistance gene product P-glycoprotein in normal human tissues. Proc Natl Acad Sci U S A 84, 7735–7738 (1987).

6. Tanigawara, Y. Role of P-glycoprotein in drug disposition. Ther Drug Monit 22, 137–140 (2000).

7. Kartner, N., Evernden-Porelle, D., Bradley, G. & Ling, V. Detection of P-glycoprotein in multidrug-resistant cell lines by monoclonal antibodies. Nature 316, 820–823 (1985).

8. Fung, K. L. & Gottesman, M. M. A synonymous polymorphism in a common MDR1 (ABCB1) haplotype shapes protein function. Biochim Biophys Acta 1794, 860–871 (2009).

9. Wang, L.-H., Song, Y.-B., Zheng, W.-L., Jiang, L. & Ma, W.-L. The association between polymorphisms in the MDR1 gene and risk of cancer: A systematic review and pooled analysis of 52 case-control studies. Cancer Cell Int 13, 46 (2013).

10. Panczyk, M. et al. ABCB1 gene polymorphisms and haplotype analysis in colorectal cancer. Int J Colorectal Dis 24, 895–905 (2009).

11. Sherry, S. T. et al. dbSNP: the NCBI database of genetic variation. Nucleic Acids Res 29, 308–311 (2001).

12. Spooner, W. et al. Haplosaurus computes protein haplotypes for use in precision drug design. Nat Commun 9, 4128 (2018).

13. Wang, S. Y. et al. A synonymous mutation in IGF-1 impacts the transcription and translation process of gene expression. Mol Ther Nucleic Acids 26, 1446–1465 (2021).

14. Tarrant, D. & von der Haar, T. Synonymous codons, ribosome speed, and eukaryotic gene expression regulation. Cell Mol Life Sci 71, 4195–4206 (2014).

15. Walsh, I. M., Bowman, M. A., Soto Santarriaga, I. F., Rodriguez, A. & Clark, P. L. Synonymous codon substitutions perturb cotranslational protein folding in vivo and impair cell fitness. Proc Natl Acad Sci U S A 117, 3528–3534 (2020).

16. Gu, W., Wang, X., Zhai, C., Xie, X. & Zhou, T. Selection on synonymous sites for increased accessibility around miRNA binding sites in plants. Mol Biol Evol 29, 3037– 3044 (2012).

17. Mueller, W. F., Larsen, L. S. Z., Garibaldi, A., Hatfield, G. W. & Hertel, K. J. The Silent Sway of Splicing by Synonymous Substitutions. J Biol Chem 290, 27700–27711 (2015).

18. Robey, R. W. et al. Revisiting the role of ABC transporters in multidrug-resistant cancer. Nat Rev Cancer 18, 452–464 (2018).

19. He, H. et al. Association of ABCB1 polymorphisms with prognostic outcomes of anthracycline and cytarabine in Chinese patients with acute myeloid leukemia. Eur J Clin Pharmacol 71, 293–302 (2015).

20. Hemauer, S. J. et al. Modulation of human placental P-glycoprotein expression and activity by MDR1 gene polymorphisms. Biochem Pharmacol 79, 921–925 (2010).

21. Hoffmeyer, S. et al. Functional polymorphisms of the human multidrug-resistance gene: multiple sequence variations and correlation of one allele with P-glycoprotein expression and activity in vivo. Proc Natl Acad Sci U S A 97, 3473–3478 (2000).

22. Song, P. et al. G2677T and C3435T genotype and haplotype are associated with hepatic ABCB1 (MDR1) expression. J Clin Pharmacol 46, 373–379 (2006).

23. Pang, L. et al. ATP-Binding Cassette Genes Genotype and Expression: A Potential Association with Pancreatic Cancer Development and Chemoresistance? Gastroenterol Res Pract 2014, 414931 (2014).

24. Kimchi-Sarfaty, C. et al. A ‘silent’ polymorphism in the MDR1 gene changes substrate specificity. Science 315, 525—528 (2007).

25. Kroetz, D. L. et al. Sequence diversity and haplotype structure in the human ABCB1 (MDR1, multidrug resistance transporter) gene. Pharmacogenetics 13, 481–494 (2003).

26. Gow, J. M., Hodges, L. M., Chinn, L. W. & Kroetz, D. L. Substrate-dependent effects of human ABCB1 coding polymorphisms. J Pharmacol Exp Ther 325, 435–442 (2008).

27. Hung, C.-C., Chen, C.-C., Lin, C.-J. & Liou, H.-H. Functional evaluation of polymorphisms in the human ABCB1 gene and the impact on clinical responses of antiepileptic drugs. Pharmacogenet Genomics 18, 390–402 (2008).

28. Salama, N. N., Yang, Z., Bui, T. & Ho, R. J. Y. MDR1 haplotypes significantly minimize intracellular uptake and transcellular P-gp substrate transport in recombinant LLC-PK1 cells. J Pharm Sci 95, 2293–2308 (2006).

29. Fung, K. L. et al. MDR1 synonymous polymorphisms alter transporter specificity and protein stability in a stable epithelial monolayer. Cancer Res 74, 598–608 (2014).

30. Ni, L.-N. et al. Multidrug resistance gene (MDR1) polymorphisms correlate with imatinib response in chronic myeloid leukemia. Med Oncol 28, 265–269 (2011).

31. Lu, Y. et al. Host genetic variants of ABCB1 and IL15 influence treatment outcome in paediatric acute lymphoblastic leukaemia. Br J Cancer 110, 1673–1680 (2014).

32. Zheng, Q. et al. ABCB1 polymorphisms predict imatinib response in chronic myeloid leukemia patients: a systematic review and meta-analysis. Pharmacogenomics J 15, 127– 134 (2015).

33. Chu, Y.-H. et al. Association of ABCB1 and FLT3 Polymorphisms with Toxicities and Survival in Asian Patients Receiving Sunitinib for Renal Cell Carcinoma. PLoS One 10, e0134102 (2015).

34. Munisamy, M. et al. Pharmacogenetics of ATP binding cassette transporter MDR1(1236C>T) gene polymorphism with glioma patients receiving Temozolomide-based chemoradiation therapy in Indian population. Pharmacogenomics J 21, 262–272 (2021).

35. Li, J. Z., Tian, Z. Q., Jiang, S. N. & Feng, T. Effect of variation of ABCB1 and GSTP1 on osteosarcoma survival after chemotherapy. Genet Mol Res 13, 3186–3192 (2014).

36. Olarte Carrillo, I., et al. High expression levels and the C3435T SNP of the ABCB1 gene are associated with lower survival in adult patients with acute myeloblastic leukemia in Mexico City. BMC Med Genomics 14, 251 (2021).

37. Balcerczak, E. et al. ABCB1/MDR1 gene polymorphisms as a prognostic factor in colorectal cancer. Int J Colorectal Dis 25, 1167–1176 (2010).

38. Caronia, D. et al. Effect of ABCB1 and ABCC3 polymorphisms on osteosarcoma survival after chemotherapy: a pharmacogenetic study. PLoS One 6, e26091 (2011).

39. Wu, H. et al. Roles of ABCB1 gene polymorphisms and haplotype in susceptibility to breast carcinoma risk and clinical outcomes. J Cancer Res Clin Oncol 138, 1449–1462 (2012).

40. Knez, L. et al. Predictive value of ABCB1 polymorphisms G2677T/A, C3435T, and their haplotype in small cell lung cancer patients treated with chemotherapy. J Cancer Res Clin Oncol 138, 1551–1560 (2012).

41. Vivona, D. et al. ABCB1 haplotypes are associated with P-gp activity and affect a major molecular response in chronic myeloid leukemia patients treated with a standard dose of imatinib. Oncol Lett 7, 1313–1319 (2014).

42. Li, W. et al. ABCB1 3435TT and ABCG2 421CC genotypes were significantly associated with longer progression-free survival in Chinese breast cancer patients. Oncotarget 8, 111041–111052 (2017).

43. Gregers, J. et al. Polymorphisms in the ABCB1 gene and effect on outcome and toxicity in childhood acute lymphoblastic leukemia. Pharmacogenomics J 15, 372–379 (2015).

44. Xiaohui, S. et al. Effect of ABCB1 polymorphism on the clinical outcome of osteosarcoma patients after receiving chemotherapy. Pak J Med Sci 30, 886–890 (2014).

45. Liu, S. et al. Predictive potential of ABCB1, ABCC3, and GSTP1 gene polymorphisms on osteosarcoma survival after chemotherapy. Tumour Biol 35, 9897–9904 (2014).

46. Zmorzynski, S. et al. The Relationship of ABCB1/MDR1 and CYP1A1 Variants with the Risk of Disease Development and Shortening of Overall Survival in Patients with Multiple Myeloma. J Clin Med 10, (2021).

47. Chen, Q. et al. Prognostic Value of Two Polymorphisms, rs1045642 and rs1128503, in ABCB1 Following Taxane-based Chemotherapy: A Meta-Analysis. Asian Pac J Cancer Prev 22, 3–10 (2021).

48. Graudejus, O., Wong, R., Varghese, N., Wagner, S. & Morrison, B. Bridging the gap between in vivo and in vitro research: Reproducing in vitro the mechanical and electrical environment of cells in vivo. Front Cell Neurosci 12, (2018).

49. Cunningham, F. et al. Ensembl 2022. Nucleic Acids Res 50, D988–D995 (2022).

50. Wang, M. et al. PaxDb, a Database of Protein Abundance Averages Across All Three Domains of Life. Mol Cell Proteomics 11, 492–500 (2012).

51. Karson, M. Handbook of Methods of Applied Statistics. Volume I: Techniques of Computation Descriptive Methods, and Statistical Inference. Volume II: Planning of Surveys and Experiments. I. M. Chakravarti, R. G. Laha, and J. Roy, New York, John Wiley; 1967, $9.00. J Am Stat Assoc 63, 1047–1049 (1968).

52. Avsec, Ž., et al. Effective gene expression prediction from sequence by integrating long-range interactions. Nat Methods 18, 1196–1203 (2021).

53. Jaganathan, K. et al. Predicting Splicing from Primary Sequence with Deep Learning. Cell 176, (2019).

54. Hofacker, I. et al. Automatic Detection of Conserved RNA Structure Elements in Complete RNA Virus Genomes. Nucleic Acids Res 26, 3825–3836 (1998).

55. Sharp, P. & Li, W.-H. The codon Adaptation Index—A measure of directional synonymous codon usage bias, and its potential applications. Nucleic Acids Res 15, 1281– 1295 (1987).

56. Nakamura, Y., Gojobori, T. & Ikemura, T. Codon usage tabulated from international DNA sequence databases: status for the year 2000. Nucleic Acids Res 28, 292 (2000).

57. Hernandez-Alias, X., Benisty, H., Schaefer, M. H. & Serrano, L. Translational adaptation of human viruses to the tissues they infect. Cell Rep 34, 108872 (2021).

58. Sabi, R. & Tuller, T. Modelling the Efficiency of Codon–tRNA Interactions Based on Codon Usage Bias. DNA Research 21, 511–526 (2014).

59. Yu, C.-H. et al. Codon Usage Influences the Local Rate of Translation Elongation to Regulate Co-translational Protein Folding. Mol Cell 59, 744–754 (2015).

60. Sievers, F. et al. Fast, scalable generation of high-quality protein multiple sequence alignments using Clustal Omega. Mol Syst Biol 7, 539 (2011).

61. Suyama, M., Torrents, D. & Bork, P. Suyama M, Torrents D, Bork P.. PAL2NAL: robust conversion of protein sequence alignments into the corresponding codon alignments. Nucleic Acids Res 34: W609–W612. *Nucleic Acids Res* **34**, W609-12 (2006).

62. Kaplan, E. L. & Meier, P. Nonparametric Estimation from Incomplete Observations. in Breakthroughs in Statistics: Methodology and Distribution (eds. Kotz, S. & Johnson, N. L.) 319–337 (Springer New York, 1992). doi:10.1007/978-1-4612-4380-9_25.

63. Mantel, N. Evaluation of survival data and two new rank order statistics arising in its consideration. Cancer Chemother Rep 50, 163–170 (1966).

64. Karollus, A., Mauermeier, T. & Gagneur, J. Current sequence-based models capture gene expression determinants in promoters but mostly ignore distal enhancers. bioRxiv 2022.09.15.508087 (2022) doi:10.1101/2022.09.15.508087.

65. Sasse, A. et al. How far are we from personalized gene expression prediction using sequence-to-expression deep neural networks? bioRxiv 2023.03.16.532969 (2023) doi:10.1101/2023.03.16.532969.

66. Kudla, G., Murray, A. W., Tollervey, D. & Plotkin, J. B. Coding-sequence determinants of gene expression in Escherichia coli. Science 324, 255–258 (2009).

67. Futcher, B., Latter, G. I., Monardo, P., McLaughlin, C. S. & Garrels, J. I. A Sampling of the Yeast Proteome. Mol Cell Biol 19, 7357–7368 (1999).

68. dos Reis, M., Wernisch, L. & Savva, R. Unexpected correlations between gene expression and codon usage bias from microarray data for the whole Escherichia coli K-12 genome. Nucleic Acids Res 31, 6976–6985 (2003).

69. Waldman, Y. Y., Tuller, T., Shlomi, T., Sharan, R. & Ruppin, E. Translation efficiency in humans: tissue specificity, global optimization and differences between developmental stages. Nucleic Acids Res 38, 2964–2974 (2010).

70. Hardesty, B., Tsalkova, T. & Kramer, G. Co-translational folding. Curr Opin Struct Biol 9, 111–114 (1999).

71. Binkhathlan, Z. & Lavasanifar, A. P-glycoprotein Inhibition as a Therapeutic Approach for Overcoming Multidrug Resistance in Cancer: Current Status and Future Perspectives. Curr Cancer Drug Targets 13, (2013).

72. Johnatty, S. E. et al. ABCB1 (MDR1) polymorphisms and ovarian cancer progression and survival: A comprehensive analysis from the Ovarian Cancer Association Consortium and The Cancer Genome Atlas. Gynecol Oncol 131, 8–14 (2013).

