## Supplementary Figures for "Revisiting the effects of MDR1 Variants using computational approaches"

***Figure S1: Main phases of gene expression.*** *Transcription: A region of the double helix of the DNA is unwound and a pre-mRNA sequence is transcribed using one of the DNA strands as templet. Splicing: The pre-mRNA is edited and introns are removed from the it. Additionally, a 5' cap and 3' poly-A tail are added to the mRNA. Translation: The ribosome synthesizes a protein according to the mRNA templet. During the translation process the nascent protein initiates the formation of secondary and tertiary structures. mRNA degradation: mRNAs undergo degradation and are broken to smaller fragments, mainly by ribonucleases.*

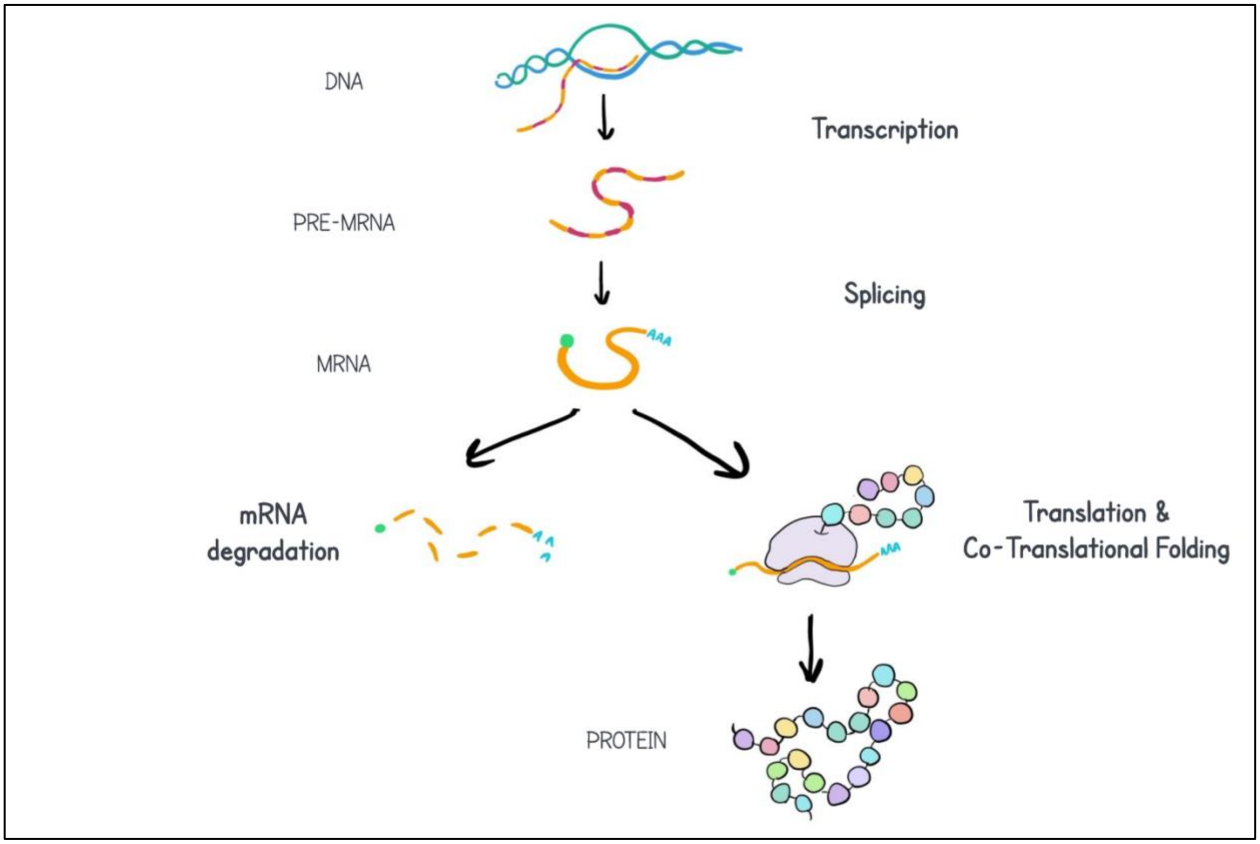


***Figure S2: MDR1 expression levels of carriers vs. non-carriers of TCGA mutations that are highly correlated with T1236C (present in the genomes of over 75% of T1236C positive patients).***
*Comparison of the mean MDR1 expression of the carriers group (vertical line) to the mean MDR1 expression levels of 100,000 groups of randomly chosen non-carriers (distribution). Size of the carriers' group and non-carriers' groups are the same.*

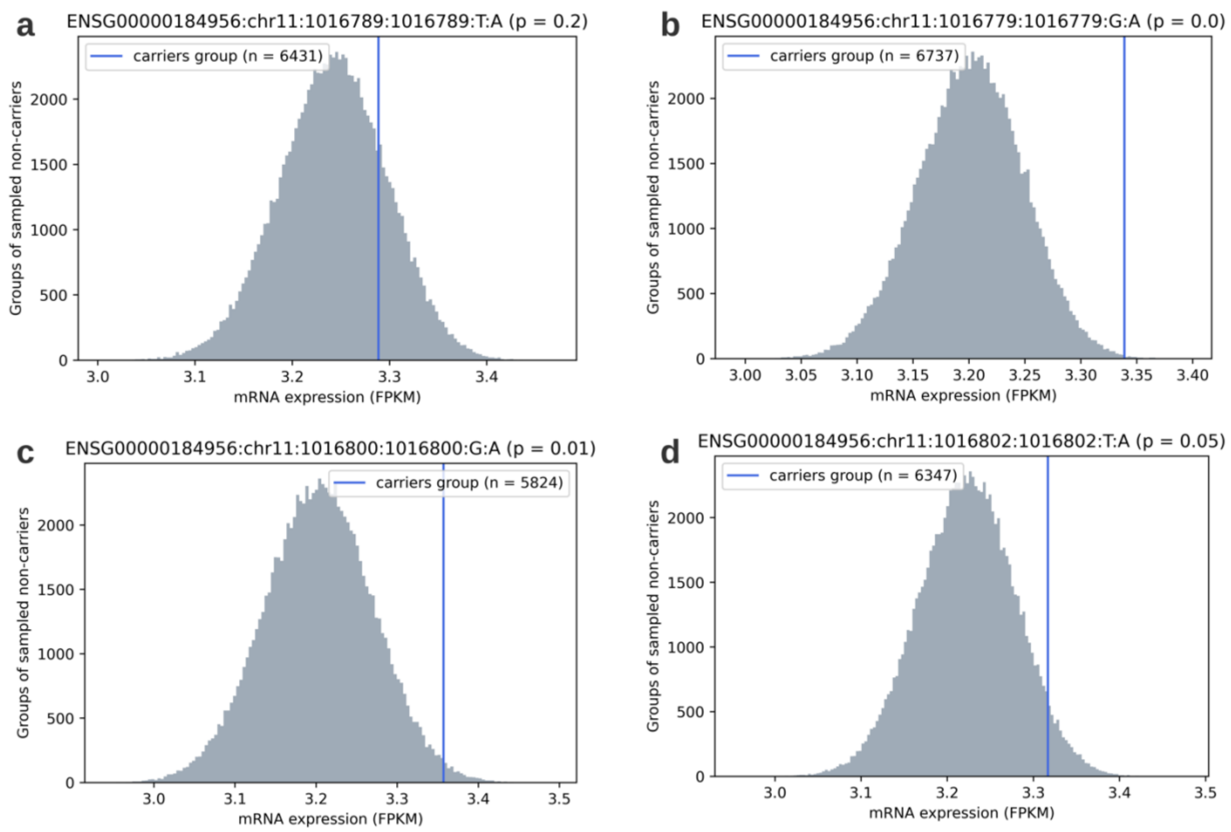


***Figure S3: MDR1 expression levels of carriers vs. non-carriers of the combinations of the three variants, from the TCGA database.*** *a**:** T1236C & T2677G ; b: T1236C & T3435C ; c: T2677G & T3435C;* ***.*** *d**:** T1236C, T2677G & T3435C. Comparison of the mean MDR1 expression of the carriers group (vertical line) to the mean MDR1 expression levels of 100,000 groups of randomly chosen non-carriers (distribution). Size of the carriers' group and non-carriers' groups are the same.*

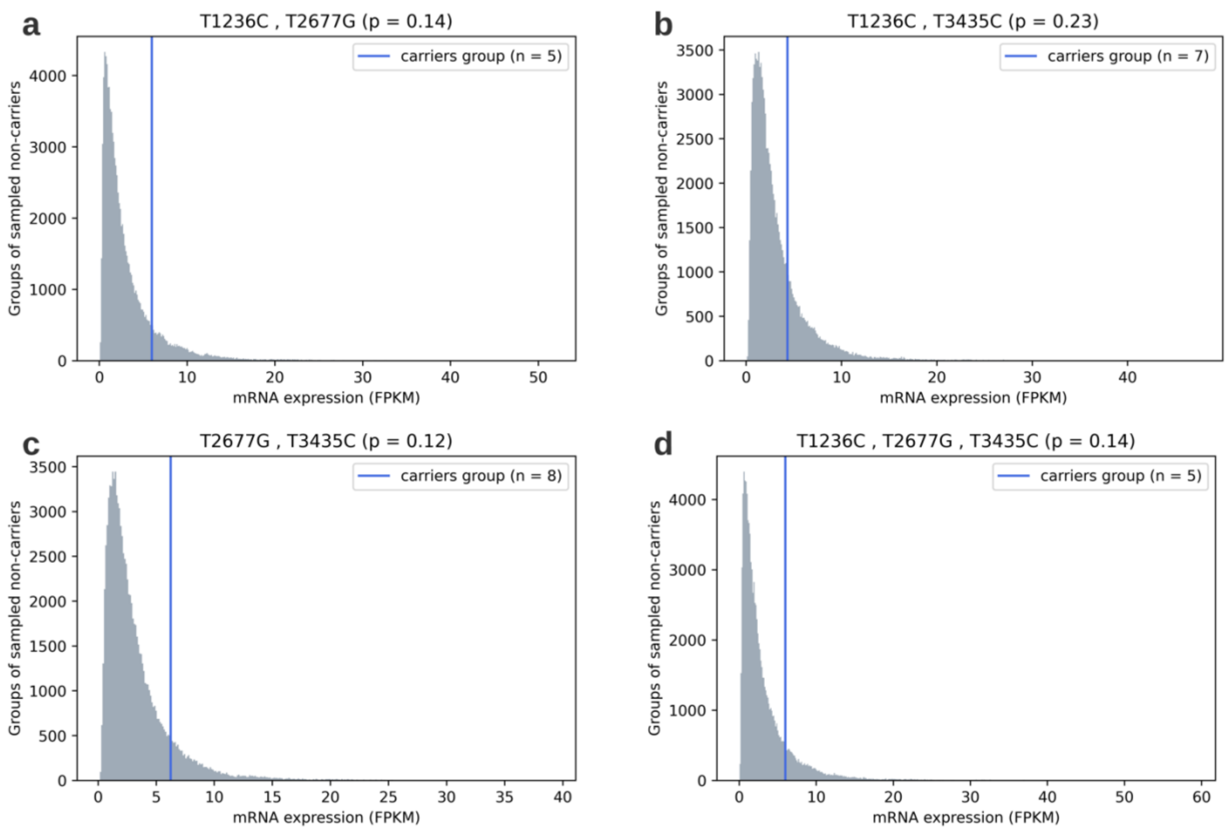


***Figure S4: MFE profile of the unmutated CDS sequence of MDR1.*** *The positions of the variants are denoted in blue (1236), orange (2677) and green (3435). The percentile of the MFE score of the positions of the variants (when unmutated) is denoted in the legend.*

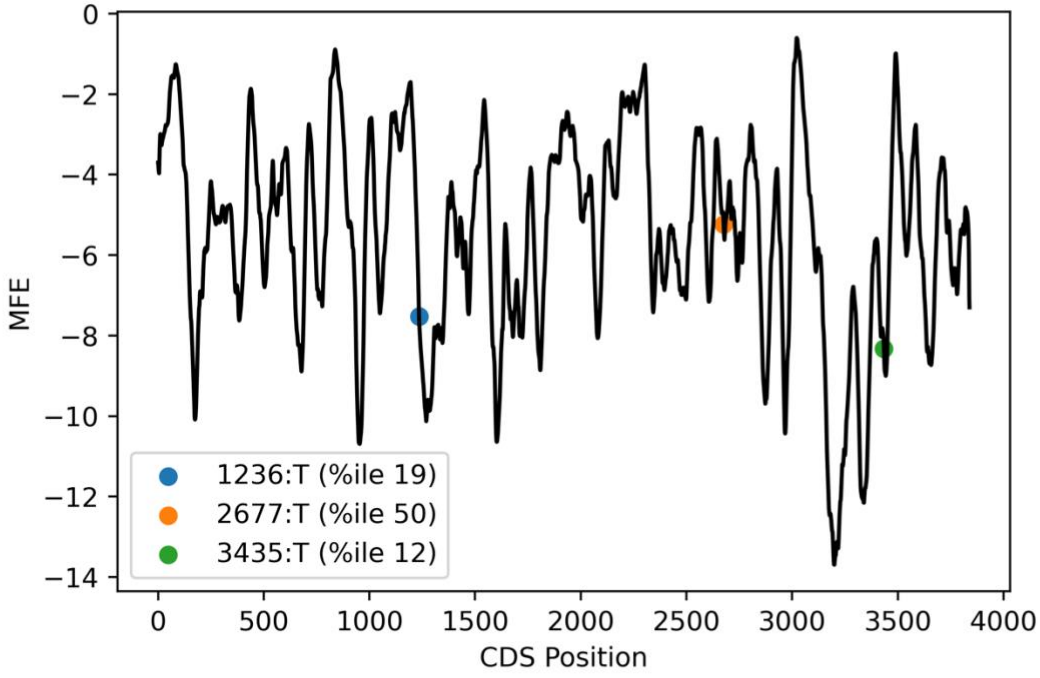

***Figure S5: Effect of the three variants on tAI in the kidney tissue.*** *a: T1236C ; b: T2677G ; c: T3435C .x axis: the amino-acid position in the protein sequence; y axis: the tAI score. The vertical line indicates the position of the variant. Black dots indicate the (unchanged) tAI scores in the vicinity of the variant and the dots on the vertical line indicate the tAI score of the mutated position, before (green) and after (pink).*

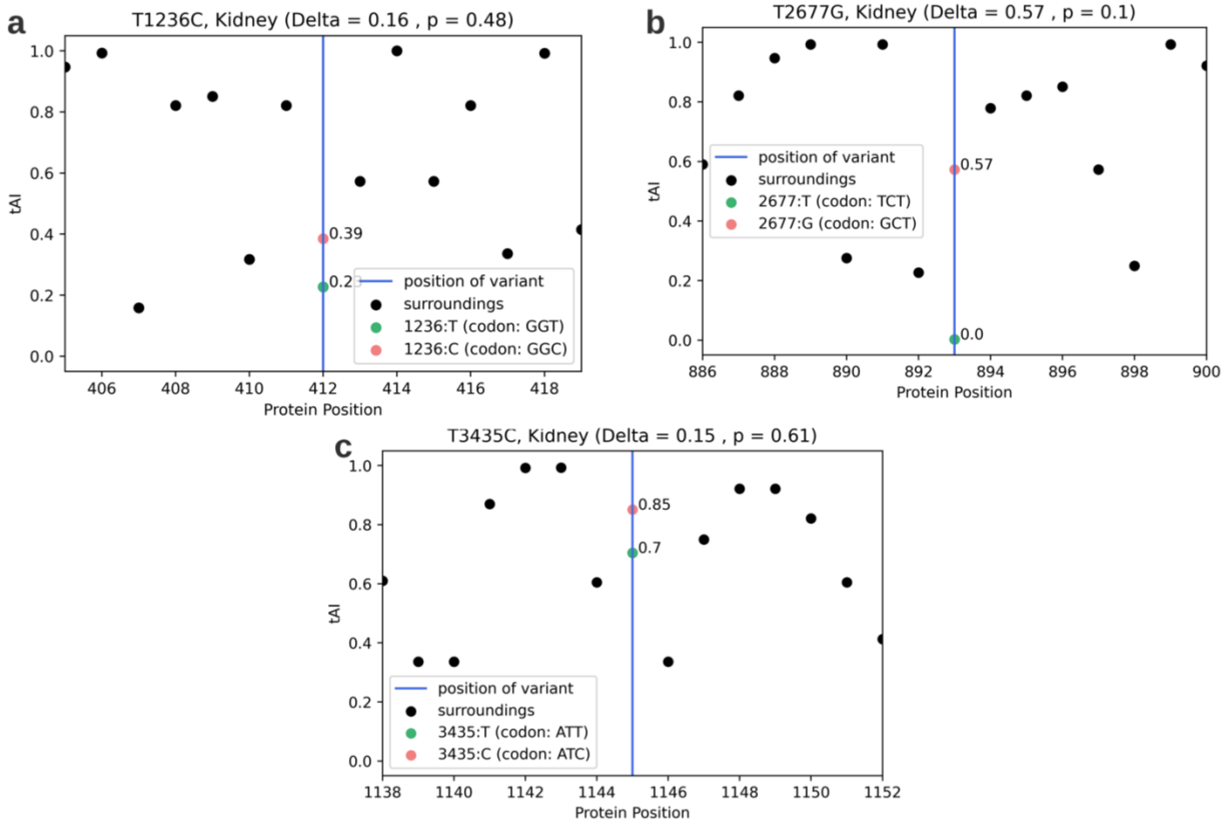


***Figure S6: Effect of the three variants on tAI in the colon tissue.*** *a: T1236C ; b: T2677G ; c: T3435C .x axis: the amino-acid position in the protein sequence; y axis: the tAI score. The vertical line indicates the position of the variant. Black dots indicate the (unchanged) tAI scores in the vicinity of the variant and the dots on the vertical line indicate the tAI score of the mutated position, before (green) and after (pink).*

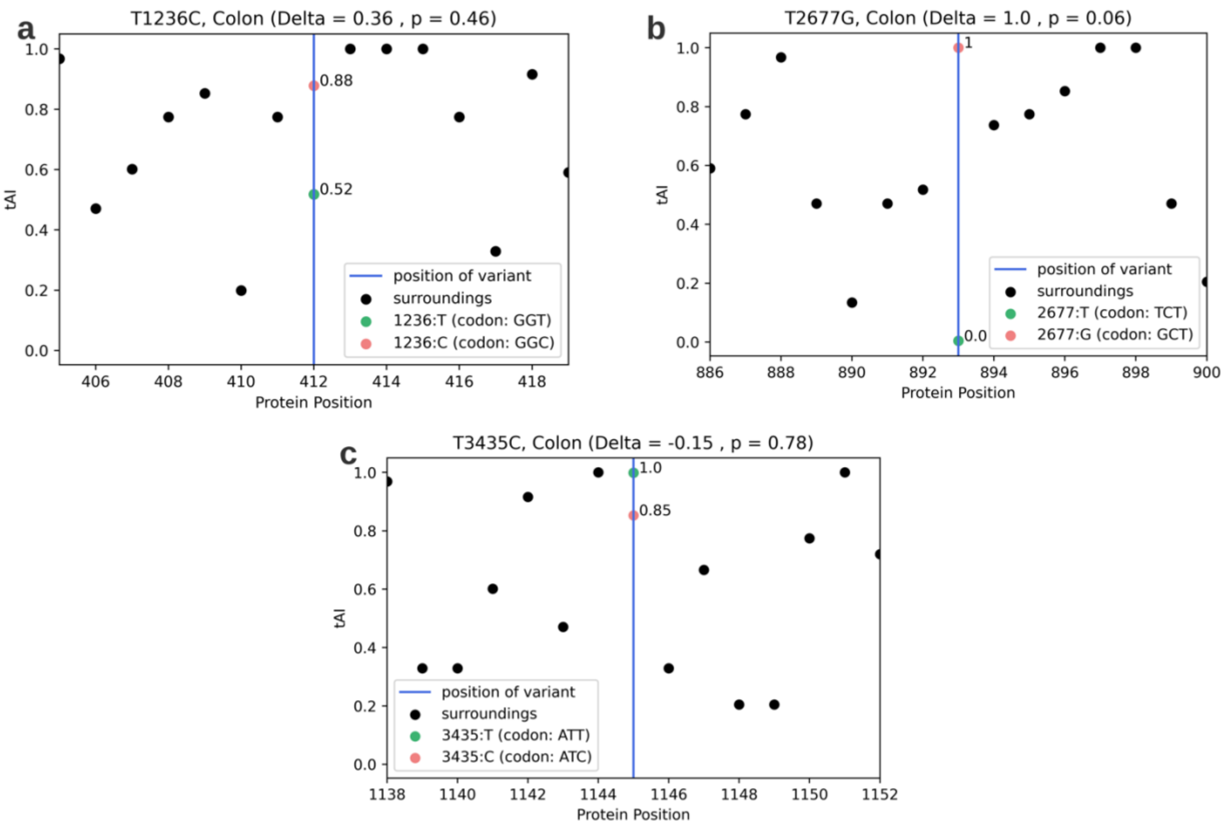


***Figure S7: Effect of the three variants on tAI in the brain tissue.*** *a: T1236C ; b: T2677G ; c: T3435C .x axis: the amino-acid position in the protein sequence*

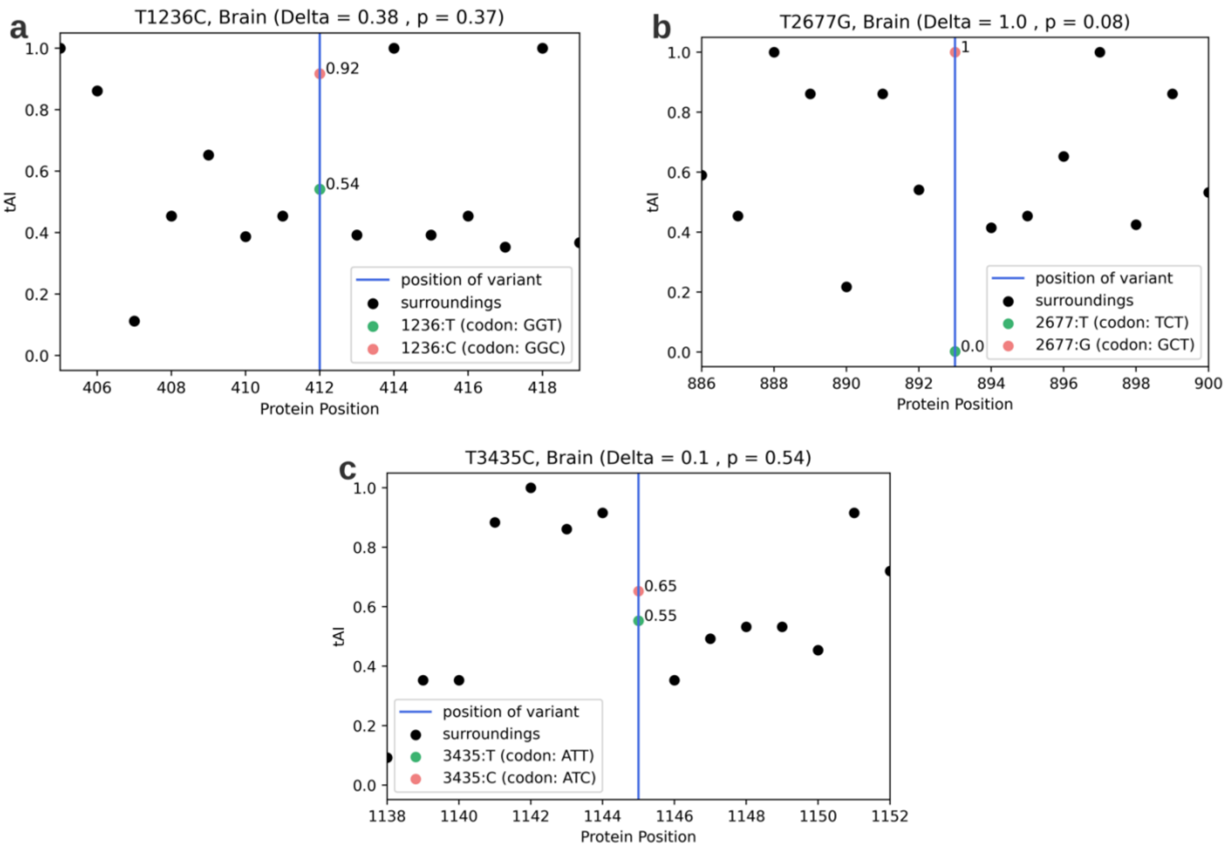


*; y axis: the tAI score. The vertical line indicates the position of the variant. Black dots indicate the (unchanged) tAI scores in the vicinity of the variant and the dots on the vertical line indicate the tAI score of the mutated position, before (green) and after (pink).*

***Figure S8: Kaplan-Meier survival curves of the carriers and non-carriers of the combinations of the three variants, from the TCGA database.*** ***a:*** *T1236C & T2677G ; b: T1236C & T3435C ; c: T2677G & T3435C;* ***.*** *d**:** T1236C, T2677G & T3435C.*

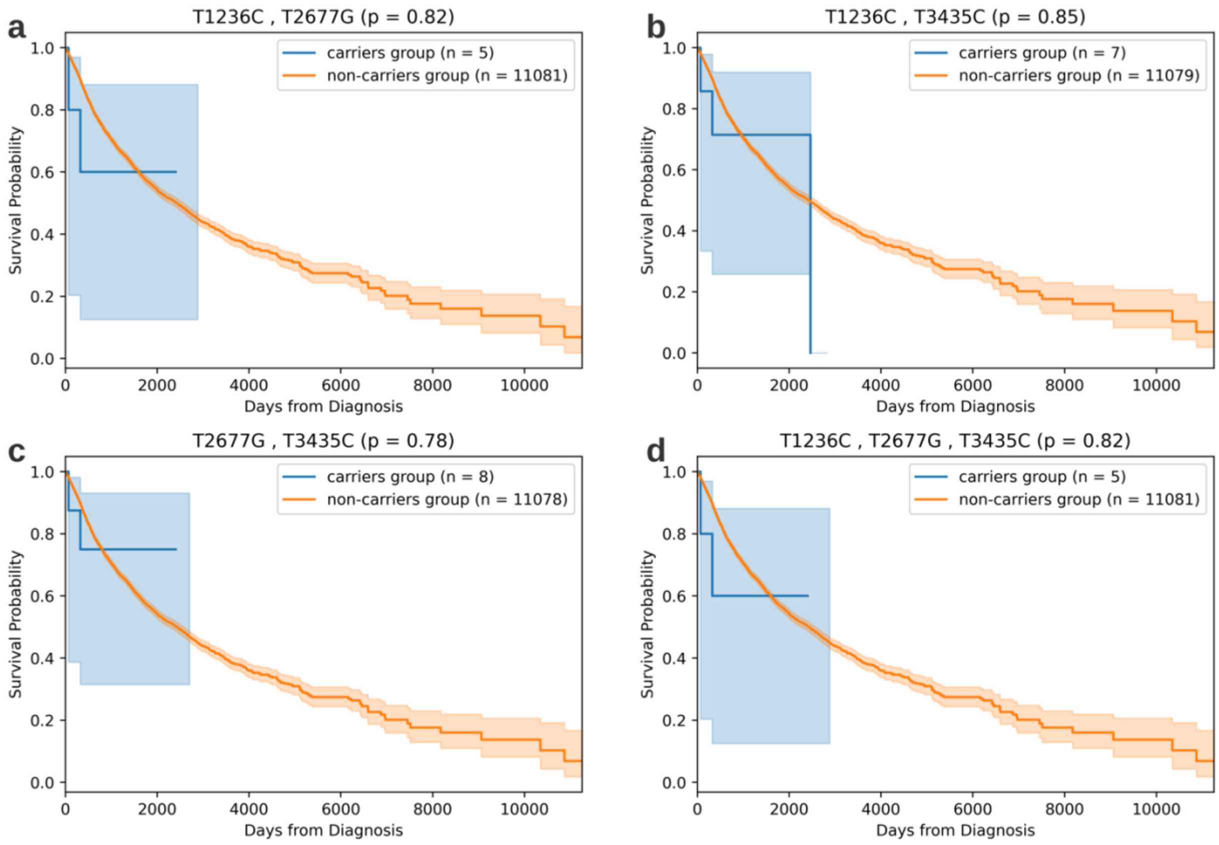


***Figure S9: Kaplan-Meier survival curves of carriers vs. non-carriers of TCGA mutations that are highly correlated with T1236C (present in the genomes of over 75% of T1236C positive patients).***

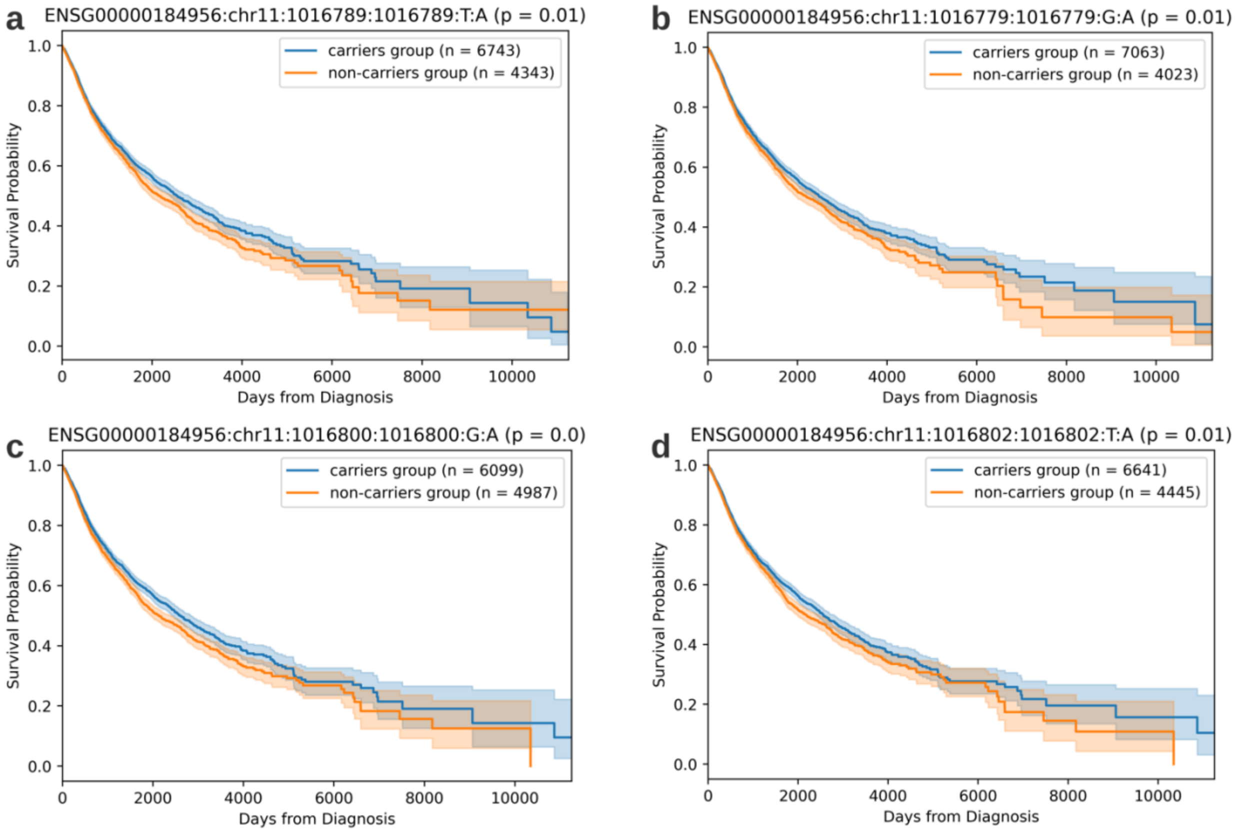


***Figure S10: Effect of the three variants on survivability of patients that underwent chemotherapy.*** *a: T1236C; b: T2677G; c: T3435C. Comparison of the Kaplan-Meier survival curves of the carriers' group and the non-carriers group of the three variants, excluding patients that have not received chemotherapy treatments.*

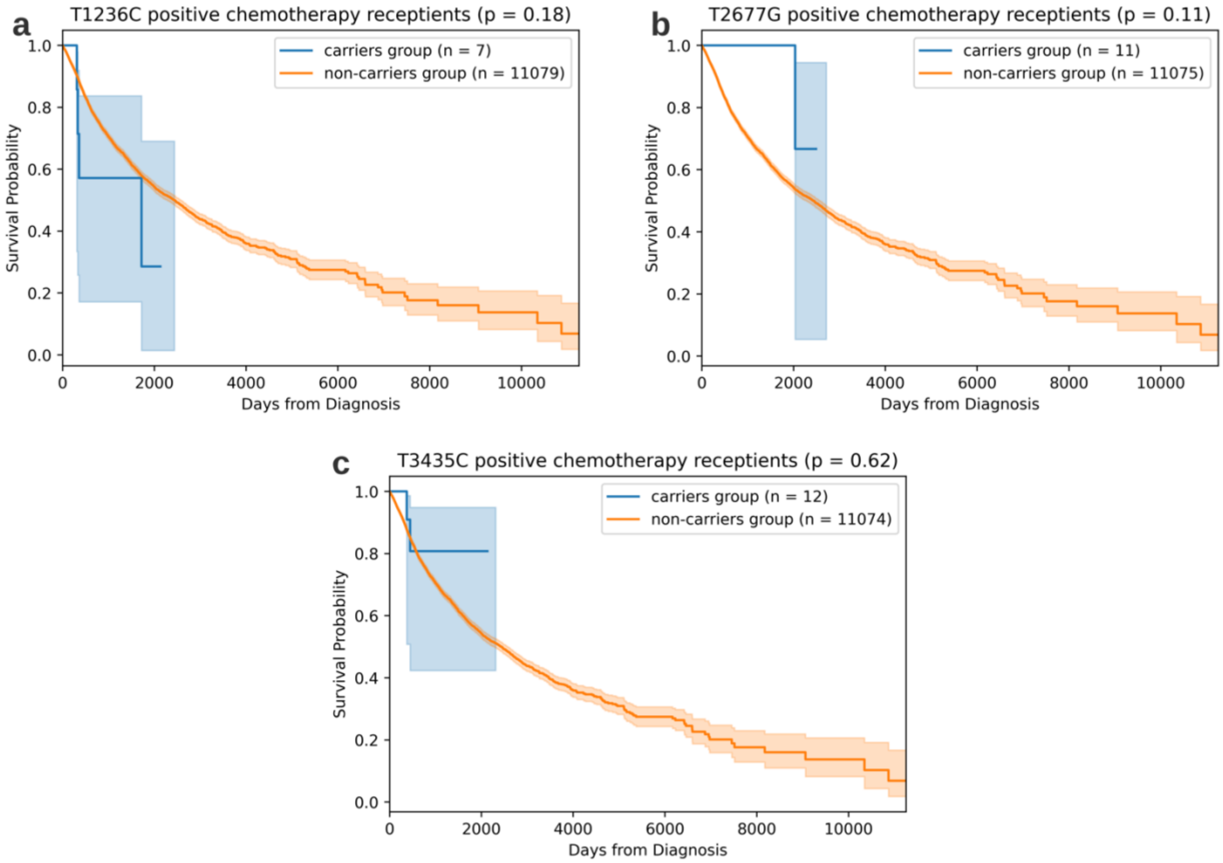
a
T1236C positive chemotherapy receptients (p = 0.18)
carriers group (n = 7)
non-carriers group (n = 11079)
Survival Probability
Days from Diagnosis
b
T2677G positive chemotherapy receptients (p = 0.11)
carriers group (n = 11)
non-carriers group (n = 11075)
Survival Probability
Days from Diagnosis
c
T3435C positive chemotherapy receptients (p = 0.62)
carriers group (n = 12)
non-carriers group (n = 11074)
Survival Probability
Days from Diagnosis
